# Accounting for the Role of Asymptomatic Patients in Understanding the Dynamics of the COVID-19 Pandemic: A Case Study from Singapore

**DOI:** 10.1101/2021.07.21.21260919

**Authors:** Liew Fu Teck, Palash Ghosh, Bibhas Chakraborty

## Abstract

**Objectives:** To forecast the true growth of COVID-19 cases in Singapore after accounting for asymptomatic infections, we study and make modifications to the SEIR (Susceptible-Exposed-Infected-Recovered) epidemiological model by incorporating hospitalization dynamics and the presence of asymptomatic cases. We then compare the simulation results of our three epidemiological models of interest against the daily reported COVID-19 case counts across the time period from 23^rd^ January to 6^th^ April 2020. Finally, we compare and evaluate on the performance and accuracy of the aforementioned models’ simulations.

**Methods:** Three epidemiological models are used to forecast the true growth of COVID-19 case counts by accounting for asymptomatic infections in Singapore. They are the exponential model, SEIR model with hospitalization dynamics (SEIHRD), and the SEIHRD model with inclusion of asymptomatic cases (SEAIHRD).

**Results:** Simulation results of all three models reflect underestimation of COVID-19 cases in Singapore during the early stages of the pandemic. At a 40% asymptomatic proportion, we report basic reproduction number *R*_0_ = 3.28 and 3.74 under the SEIHRD and SEAIHRD models respectively. At a 60% asymptomatic proportion, we report *R*_0_ = 3.48 and 3.96 under the SEIHRD and SEAIHRD models respectively.

**Conclusions:** Based on the results of different simulation scenarios, we are highly confident that the number of COVID-19 cases in Singapore was underestimated during the early stages of the pandemic. This is supported by the exponential increase of COVID-19 cases in Singapore as the pandemic developed.

## 1 Introduction

The world has been thrown into a global health crisis in 2020 due to the novel coronavirus, resulting in the disease commonly known as COVID-19. The first case of COVID-19 was detected in Wuhan, China in December 2019 and we have since seen the virus spread to more than 179 million cases and 3.88 million deaths worldwide as of June 2021 (Our World in Data, 2021). The World Health Organization (WHO) declared COVID-19 as a global pandemic on 11 March 2020 (The Straits Times, 2020) and countries all around the world subsequently took proactive precautionary measures such as temporary travel restrictions, social distancing measures and nation-wide lockdowns to minimise the gathering of crowds in close proximity over extended durations (Ministry of Health (MOH), 2020).

Despite the undertaking of these precautionary measures, there have been multiple countries and regions facing the risk of successive waves of COVID-19, such as Korea (BBC News, 2020) and UK (Ng, 2020). One widely recognised root cause behind the successive COVID-19 waves is the increasing number of asymptomatic cases that form COVID-19 clusters. These individuals show little to no symptoms but remain just as infectious as symptomatic patients, and close contacts of these individuals remain at risk of contracting the disease (Ries, 2020).

A second (and often overlooked) category consists of presymptomatic cases. This refers to the period where the individual is infected and shedding the virus but has not yet developed symptoms (Gillespie, 2021). Research has shown that this particular COVID-19 virus has a lengthy average incubation period (period between exposure to virus and symptom onset) of 5 - 6 days, and can last up to 14 days (WHO, 2020). Peak of viral shedding occurs during this incubation period and patients are the most contagious (Gillespie, 2021).

Under-testing is yet another major obstacle in keeping the pandemic under control. As discussed above, many infected COVID-19 patients who do not display symptoms (and not being tested as a result) may be spreading the disease to people in close proximity, resulting in new COVID-19 infections. These newly infected patients may also display no symptoms and are not tested, thus resulting in a vicious cycle of infection. This situation can be further exacerbated by high population densities, as in the case of USA, where there is a strong correlation between population density and the number of cumulative cases at the county level (Wong and Li, 2020).

The preceding illustration of asymptomatic and presymptomatic patients, as well as the reality of under-testing in the face of the current COVID-19 pandemic leads us to believe that current reported COVID-19 figures around the world such as the total number of COVID-19 cases and the total number of COVID-19 related deaths suffer from severe under-reporting. We hypothesize that national statistics were under-reported and the rate of transmission within the community was underestimated. Thus, we propose the usage of 3 different statistical models to simulate the spread of COVID-19 cases in Singapore.

The paper will primarily focus on the initial spread of the COVID-19 pandemic in Singapore when the first local case was reported in early 2020. As a result, the time period of the dataset used in the paper spans from 23^rd^ January 2020, up till 26^th^ January 2021. In particular, this paper will not discuss the uptick of Singapore COVID-19 cases in early 2021 - mostly imported - as a result of the Singapore government’s relaxation of border control measures.

### 1.1 COVID-19 in Singapore

Singapore is a high-income city-state in Southeast Asia. Boasting a diverse melting pot of different races and cultures, well-developed tourism industry and low crime rates, Singapore is a popular tourist destination, receiving 19.11 million international visitors in 2019 (Statista, 2021). Singapore reported its first local case of COVID-19 on 23^rd^ January 2020 - a 66 year old Chinese national who arrived in Singapore three days earlier (Yong, 2020). Following a 3-month period with a low number of daily confirmed COVID-19 cases, during which Singapore’s pandemic preparedness earned high praise from Harvard University for being the “gold standard” in case detection (Kurohi, 2020) and the WHO Director-General Tedros Adhanom Ghebreyesus for “leaving no stone unturned” (CNA, 2020), Singapore started reporting a substantial increase in the number of daily cases from mid-April onwards. To counter the spread of the virus, the government actively implemented strict preventive measures which include (but are not limited to):

– Announcement of the “circuit breaker” (comprising of stay-at-home orders and other societal restrictions with the aim of minimising transmission risks) on 7th April 2020 (MOH, 2020b)
– Mandating the wearing of masks in public (Ang and Phua, 2020)
– Travelers entering Singapore to commit to a Stay Home Notice period of 7 or 14 days (Immigration & Checkpoints Authority Singapore (ICA), 2020)

Using such precautionary measures, Singapore managed to curb the spread of the virus, as evident from a low daily COVID-19 case count (<50) in the latter half of 2020. However, there is no doubt that Singapore also suffered from under-reporting of the number of COVID-19 cases in the early stages of the pandemic. Infected but undiagnosed COVID-19 patients continued to maintain close contact with other individuals in their community, resulting in the formation of multiple COVID-19 clusters (CNA, 2021). Unfortunately, this increase in cases was not captured at the time due to our lack of understanding of the virus, as well as a testing protocol that was not yet optimized during the early months of the outbreak. Meanwhile, members of these clusters continued to infect other individuals, resulting in an exponential increase in cases. To illustrate this phenomenon, we plot the trajectory of confirmed cumulative COVID-19 cases in Singapore from 23^rd^ January 2020 to 26^th^ January 2021 (Roser et al., 2020).

From Figure 1 above, we see that Singapore’s confirmed cases remained at a steady low from 23^rd^ January 2020 when the first COVID-19 case was confirmed, up till mid-April 2020, where the number of confirmed cases increased exponentially. As discussed earlier, this sharp increase is due to a multitude of factors, including but not limited to the capturing of asymptomatic infections, as well as a revision of testing frequencies.

**Fig. 1:**
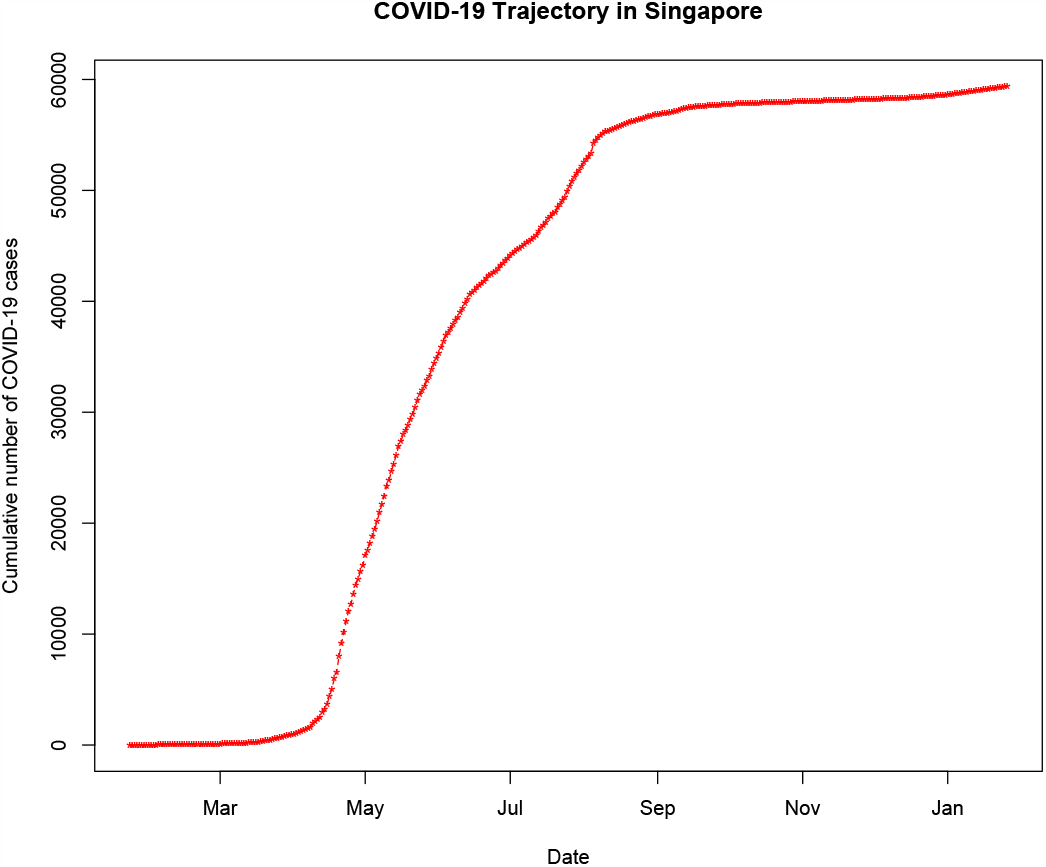
Reported cumulative COVID-19 case counts in Singapore

### 1.2 Study Objective

In this paper, we will investigate the impact of asymptomatic COVID-19 cases on the actual Singapore COVID-19 case counts during the early stages of the pandemic when measures were not in place to curb the spread of the virus - from 23^rd^ January 2020 when Singapore reported its first COVID-19 case, right up to 6^th^ April 2020, one day before the commencement of the circuit breaker (Yong, 2020). This is carried out via the construction and modification of epidemiological models, and comparing the trajectory against the reported COVID-19 case counts in Singapore.

Note that we do not make a clear distinction between the terms *presymptomatic* and *asymptomatic*, as both groups of patients do not display symptoms. We will instead classify individuals in both groups to be *asymptomatic*. This umbrella term will be used to account for both groups of COVID-19 patients in the following sections.

Discussion of the predictions in the following sections can be used to ascertain the degree of under-reporting of COVID-19 cases in Singapore as the pandemic progresses. Additionally, these figures can also serve as a guideline for health care systems to allocate finite health care resources to COVID-19 patients, and adequately prepare for successive waves of the virus or future pandemics, if necessary.

The rest of the article is organized as follows: Section 2 introduces the different infection models used and proposes modifications to the SEIR compartmental model to account for the epidemiological characteristics of COVID-19. Section 3 presents the results of the different models against the confirmed COVID-19 case counts in Singapore, while Section 4 discusses the simulation results and model limitation(s) in detail. Finally, concluding remarks are presented in Section 5.

## 2 Methodology

In this paper, we consider the use of 3 different epidemiological models. We include the use of an exponential model, which is commonly used in pandemic simulation (Chowell et al., 2016). However, the usage of any single model alone can be misleading. Hence, we will also implement 2 variations of the SEIR (Susceptible-Exposed-Infected-Recovered) model to simulate Singapore’s COVID-19 trajectory from 23^rd^ January to 6^th^ April 2020 against the trajectory of confirmed cases against Singapore’s reported COVID-19 cases across the same timeline. The SEIR model is chosen due to its analytical simplicity as compared to other mathematical models (Adiga et al., 2020), given the lack of accurate COVID-19 data. Furthermore, the differential equations used give rise to an analytical solution, allowing us to quantitatively study the impact and interactions of different compartments. Finally, one can easily modify the differential equations to suit the epidemiological characteristics of any given virus. This modeling approach would allow us to understand the severity of the pandemic from different perspectives, with the main goal to derive more accurate predictions.

### 2.1 Exponential Model

A pandemic often displays exponential growth at the initial stage. For example, the 2014-15 Ebola epidemic in West Africa displayed an exponential growth (Viboud et al., 2016). The formula for exponential growth of a variable *x* at the growth rate *r*, as time *t* goes on in discrete intervals (that is, at integer times 0, 1, 2, 3, …), is as follows:

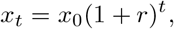

where *x*_0_ = value of *x* at time 0, *r* = growth rate and *t* = discrete time intervals (0, 1, 2, 3, …).

In our analysis, *x*_0_ is taken to be 1 (first confirmed case of COVID-19 in Singapore on 26^th^ January 2020). Simulation is carried out by taking different quantiles of the growth rates of reported cumulative COVID-19 case counts in Singapore as shown in Table 1. The growth rate at each time point *t* is calculated using the formula:

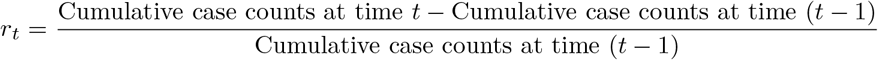

**Tab. 1:** COVID-19 case count growth rate quantiles and corresponding growth rates

| Quantile | Growth rate($r$) |
| --- | --- |
| 70% | 0.10 |
| 75% | 0.12 |
| 80% | 0.13 |
| 85% | 0.16 |
| 90% | 0.22 |

Higher quantiles of the growth rate are selected upon consideration of the skew of the growth rates, as well as the maximum growth rate being flawed due to the small number of cases in the early stages of the pandemic. This is evident from the heavily right-skewed growth rate of COVID-19 case counts in our dataset as shown in Figure 2.

**Fig. 2:**
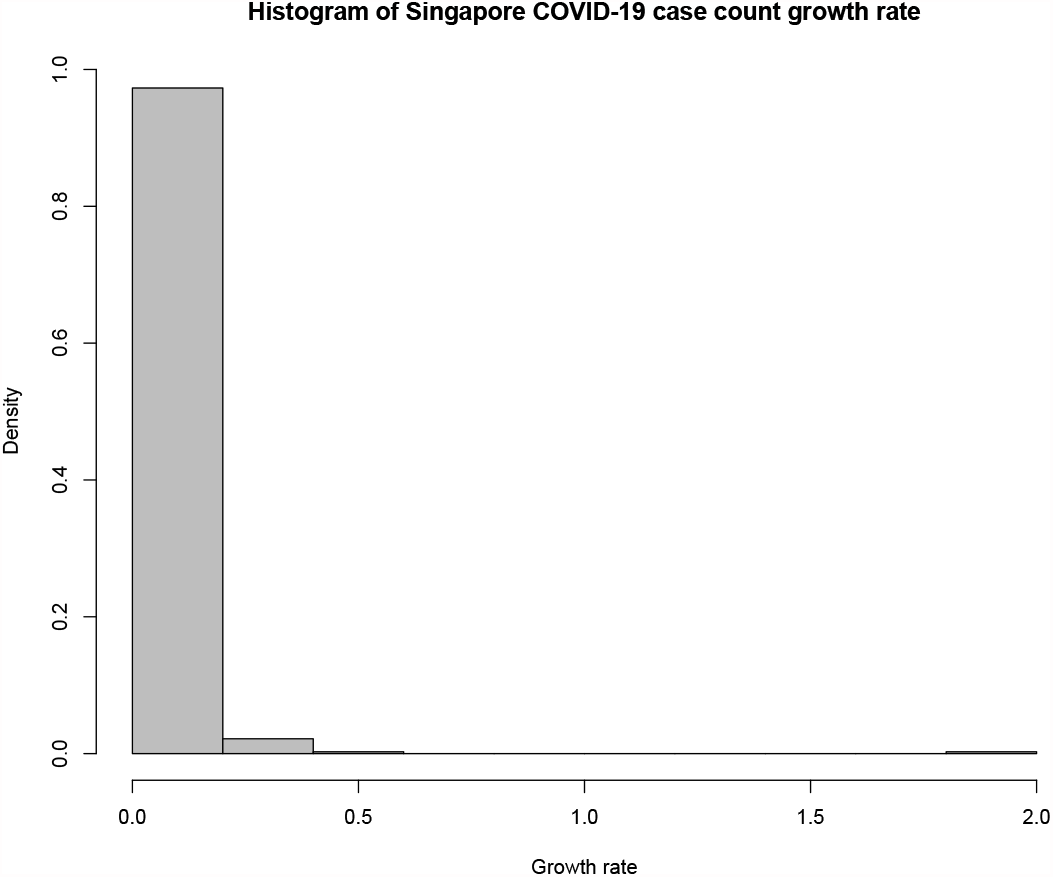
Histogram of COVID-19 case count growth rates from 23^rd^ January 2020 to 26th January 2021

**Fig. 3:**
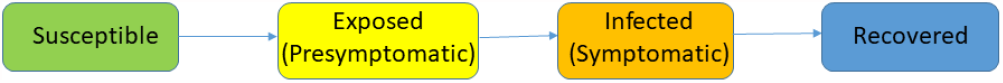
Illustration of SEIR model

### 2.2 SEIR Model

Epidemiological models are widely used to simulate the spread of infectious diseases. Within a model, each individual may progress between compartments. The SEIR (Susceptible-Exposed-Infected-Recovered) model can be used to simulate the dynamic interaction of people in 4 different conditions: susceptible (S), exposed (E), infected (I) and recovered (R) (Godio et al., 2020).

For an unmodified SEIR model, the 4 compartments consist of the following individuals:

– Susceptible(S) - Individuals at risk of contracting the virus. When a susceptible and an infectious individual come into “infectious contact”, the susceptible individual contracts the disease and transitions to the exposed (E) compartment
– Exposed(E) - Individuals who are undergoing an incubation period during which individuals are presymptomatic (i.e. infected but yet to display any symptoms)
– Infected(I) - Infectious individuals with displayed symptoms and are capable of infecting susceptible individuals
– Recovered(R) - Recovered individuals to be removed from the model (via recovery or death from virus)

### 2.3 SEIHRD Model

We propose the use of a SEIR model with hospitalization dynamics to model the COVID-19 pandemic, by incorporating the hospital (H) and death (D) compartments. These compartments are used to model hospital treatments and hospitalization periods offered to patients with COVID-19, and patients who pass away due to COVID-19 respectively.

Figure 4 gives an overview of the different compartments in this model, and how each individual can transition from one stage to the other within the Singapore context.

**Fig. 4:**
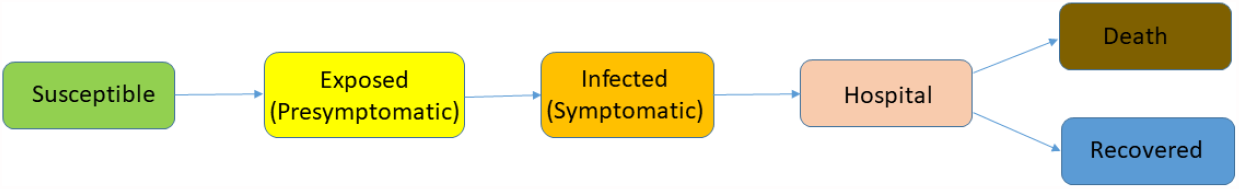
SEIR model with hospitalization and mortality dynamics

We first model the pandemic using the following differential equations and compartments from Figure 4 as a baseline model. At any predetermined time point *t, S*(*t*) denotes the susceptible population, *E*(*t*) denotes the exposed population (comprising of presymptomatic individuals), *I*(*t*) denotes the infected population, *H*(*t*) denotes the population transported to hospital for medical treatment, *R*(*t*) denotes the recovered population and *D*(*t*) denotes the dead population (due to COVID-19 complications).

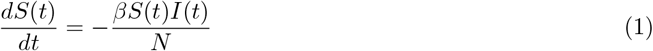

A certain proportion of susceptible individuals are exposed to the virus. These susceptible individuals are infected at a standard incidence rate (Li and Liu, 2014) of 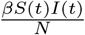

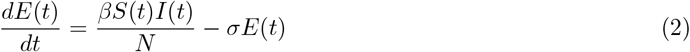

The infected individuals from equation (1) then enter the Exposed(E) compartment. COVID-19 has an incubation period ranging from anywhere between 2 to 14 days (Harvard Health Publishing, 2020) where patients are infected by the virus but do not display any symptoms yet. This compartment comprises of these presymptomatic individuals. At the end of the incubation period, we have a proportion *σ* of patients who are moved to the *I*(*t*) compartment.

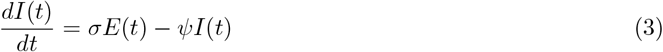

Infected-symptomatic patients leave this compartment at a rate of *ψ*, where *ψ* refers to the rate at which infected patients are sent to the hospital to receive treatment.

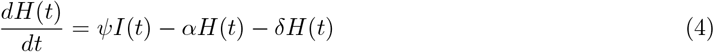

A total of *ψI*(*t*) infected patients enter the hospital to receive treatment. *α* and *δ* refer to the rate at which patients either recover from COVID-19, or die from COVID-19 complications. Hence, they leave the compartment *H*(*t*) at rates *α* and *δ* respectively.

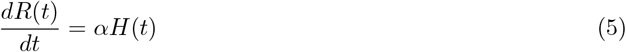

As explained above, *αH*(*t*) refers to the total number of patients recovering from COVID-19 per unit time. We define one day to be equivalent to one unit of time for the remainder of the paper.

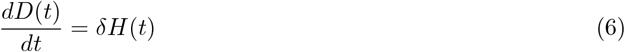

Similarly, *δH*(*t*) refers to the total number of patients passing away from COVID-19 complications per unit time.

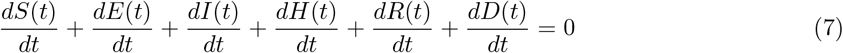

Finally, we need to check that the sum of all different partial derivatives are 0, to ensure that all compartmental changes are accounted for and total population size N is fixed (accounting for death).

The SEIHRD model is introduced as an entry point because it describes how many countries perceived and dealt with COVID-19 during the early stages of the pandemic. Nonetheless, there are certain drawbacks. Due to our lack of understanding of COVID-19 epidemiological characteristics such as the infectivity of asymptomatic COVID-19 patients, as well as an under-optimized testing protocol at the start of the outbreak, newly reported COVID-19 cases were likely symptomatic individuals who sought medical attention. However, we hypothesize that asymptomatic individuals play just as vital a role in the spread of the virus. Hence, we seek to overcome the inherent limitations of this model by proposing (and comparing against) the SEAIHRD model.

### 2.4 SEAIHRD Model

We propose further modification of the SEIHRD model in the preceding section to accurately reflect COVID-19 pandemic dynamics. To do so, we incorporate the following components:

– Segregation of I compartment into 2 sub-components: asymptomatic (A) and symptomatic (I) infections. This is done to account for virus carriers who do not show any symptoms of COVID-19 but are still capable of transmitting the COVID-19 virus to other susceptible individuals.
– Segregation of H compartment into 2 sub-components: non-ICU and ICU hospitalization. This is done to account for the differences in medical treatments offered to patients based on the severity of their symptoms.
– Inclusion of parameter *γ* to account for the quality of health care provision in Singapore (explained under Section 2.6)

Figure 5 gives an overview of the different compartments in this model, and how each individual can transition from one stage to the other within the Singapore context.

**Fig. 5:**
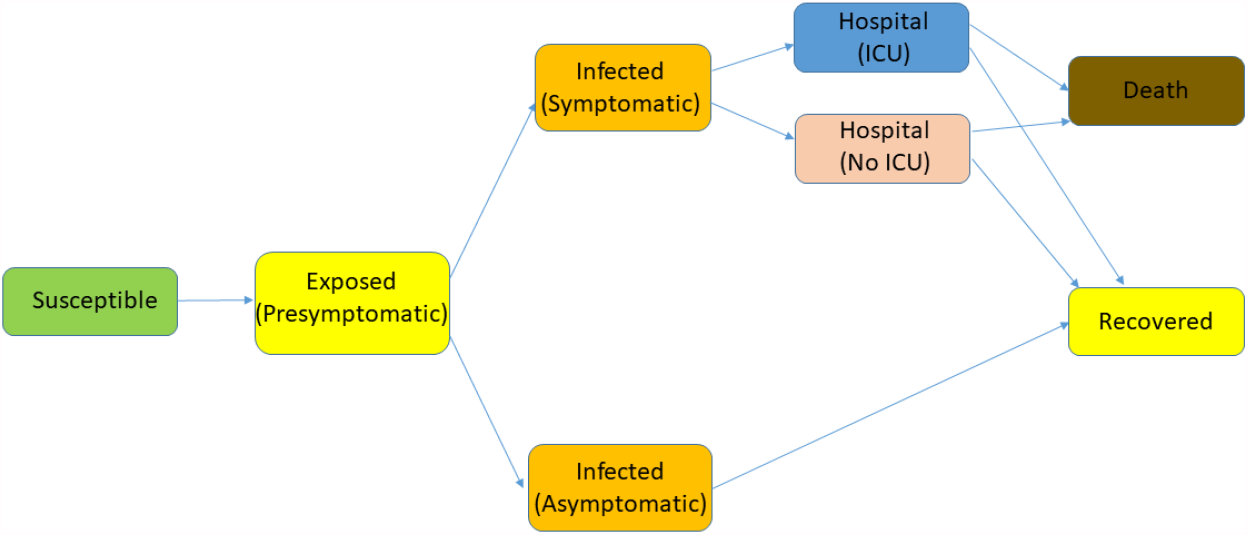
SEIHRD model with asymptomatic and presymptomatic infections

At any predetermined time point *t, S*(*t*) denotes the susceptible population, *E*(*t*) denotes the exposed population (comprising of presymptomatic individuals), *A*(*t*) denotes the asymptomatic population, *I*(*t*) denotes the infected population with symptoms, *H*_*ICU*_ (*t*) denotes the population transported to hospital ICU for emergency medical treatment, *H*_*NoICU*_(*t*) denotes the population transported to hospital (non-ICU) for general medical treatment, *R*(*t*) denotes the recovered population and *D*(*t*) denotes the dead population (due to COVID-19 complications).

We note that asymptomatic patients automatically transit to the Recovered compartment in the SEAIHRD model. In other words, this group of patients will recover from the virus without fail. Even though recent studies have shown a relation between asymptomatic COVID-19 and lung damage in the long run (Kinnear, 2020), risk of the alternative clinical outcome (Death) arising from the COVID-19 virus in the short term (as per our simulation period of 75 days) is very small (Krishnasamy et al., 2021), especially considering that the patient in question does not display any initial symptoms.

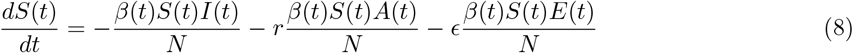

A certain proportion of susceptible individuals are exposed to the virus. These susceptible individuals are separated into 3 groups - infected-symptomatic *I*(*t*), infected-presymptomatic *E*(*t*) or infected-asymptomatic *A*(*t*). The infected-symptomatic individuals are infected at a standard incidence rate of 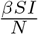, whereas the infected-asymptomatic and infected-presymptomatic individuals are infected by a standard incidence rate of 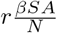 and 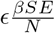 respectively (Dobrovolny, 2020). We expect constants *r* and *ϵ* to be smaller than 1, as infected-symptomatic individuals are more infectious than their asymptomatic counterparts and tend to result in a higher incidence rate of COVID-19 among their close contacts (Sayampanathan et al., 2021). We note that Figure 5 does not reflect the infectivity components from the *I*(*t*) and *A*(*t*) compartments present in Equation 8; this is because Figure 5 reflects the pathway of compartments each susceptible individual can traverse once infected with COVID-19. However, it does not reflect the “external influences” from other individuals at different stages of infection within the same population.

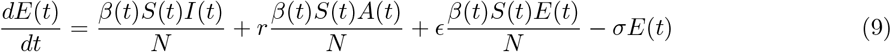

The infected individuals from equation (8) then enter the Exposed(E) compartment. COVID-19 has an incubation period ranging from anywhere between 2 to 14 days (Harvard Health Publishing, 2020) where patients are infected by the virus but do not display any symptoms yet. This compartment comprises of these presymptomatic individuals. At the end of the incubation period, a total of *σE*(*t*) patients from the *E*(*t*) compartment are moved to either the *I*(*t*) or *A*(*t*) compartment based on presence of COVID-19 symptoms.

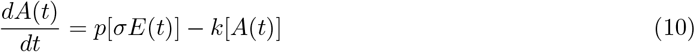

A certain proportion *p* of exposed patients who are infected but asymptomatic will enter the *A*(*t*) compartment. Asymptomatic patients are diagnosed to have fully recovered from COVID-19 after the virus leaves the body. Hence, we remove a total of *k*[*A*(*t*)] individuals from this compartment, where *k* is the rate at which asymptomatic patients recover from the virus.

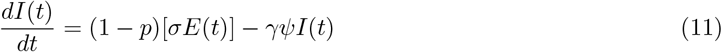

In the above, 1 - *p* refers to the proportion of exposed patients who are infected and symptomatic. Infected-symptomatic patients leave this compartment at a rate of *γψ*, where *ψ* refers to the rate at which infected patients are sent to the hospital to receive treatment. *γ* is an indicator of the country’s health care system, to be explained in detail under Section 2.6.

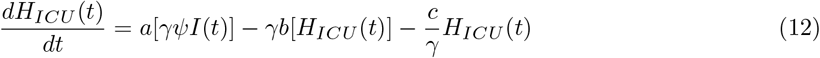

*a* refers to the proportion of patients who receive treatment in ICU. Hence, a total of *aγψI*(*t*) patients enter the hospital’s ICU. *b* and *c* refer to the rate at which ICU patients either recover from COVID-19, or die from COVID-19 complications. These ICU patients leave the *H*_*ICU*_ (*t*) compartment at rates *γb* and 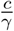 respectively.

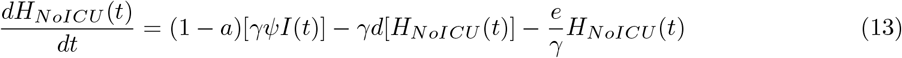

On the other hand, 1 a refers to proportion of patients receiving non-intensive care in the hospital. Hence, a total of (1 – a)*γψI*(*t*) patients receive non-intensive care within the hospital. *d* and *e* refers to the rate at which non-ICU patients either recover from COVID-19, or die from COVID-19 complications. Hence, they leave the compartment *H*_*NoICU*_ (*t*) at rates *γd* and 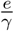 respectively.

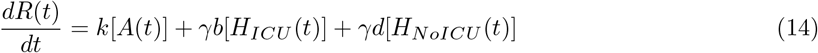

As explained above, *k*[*A*(*t*)], *γb*[*H*_*ICU*_ (*t*)] and *γd*[*H*_*NoICU*_ (*t*)] contribute to the total number of patients recovering from COVID-19 per unit time.

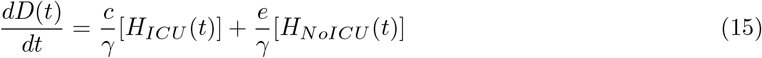

Similarly, 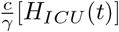 and 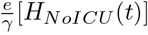 contribute to the total number of patients passing away from COVID-19 complications per unit time.

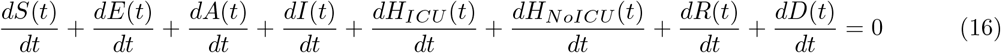

Finally, we need to check that the sum of all different partial derivatives are 0, to ensure that all compartmental changes are accounted for and total population size N is fixed (accounting for death).

### 2.5 Significance of *β*

*β* refers to the effective contact rate (Anand et al., 2021), where effective contact is defined as any kind of contact between a susceptible individual and an infected (symptomatic / asymptomatic / presymptomatic) individual such that the infected individual infects the susceptible individual with the virus. To obtain estimates of *β* in Singapore’s context, we altered the growth trajectory of Singapore’s confirmed COVID-19 cases by a multiplicative constant, based on existing studies on COVID-19 cases under-estimation.

During the early stages of the pandemic, there was no widely accepted estimate for the proportion of asymptomatic infections among COVID-19 patients. Studies carried out in different parts of the world gave rise to estimates varying from 5% all the way up to 80% (Heneghan et al., 2020). However, as our understanding of the virus grew deeper, more recent studies have given rise to a tighter band of estimates, ranging from 15.6% up to 62% (He et al., 2020; Oran and Topol, 2020; Jung et al., 2020). Based on our review of current literature, we assume 3 different degrees (20%, 40%, 60%) of under-estimation of confirmed COVID-19 cases in Singapore from the time period between 23^rd^ January 2020 and 6^th^ April 2020 and modify the number of confirmed COVID-19 cases accordingly. For each scenario of under-estimation (20%, 40%, 60%), we then optimize the value of *β* by running simulations of the 2 epidemiological models (SEIHRD, SEAIHRD) from 23^rd^ January 2020 and 6^th^ April 2020 using 1500 different *β* values and compare the growth trajectory of COVID-19 cases against the modified Singapore COVID-19 case count data. The most optimal *β* value is selected via minimization of root-mean-square error (RMSE).

We also obtain another *β* value using the same approach, but the time period of interest corresponds to when the COVID-19 case count data displayed an exponential trend from 2^nd^ May 2020 to 15^th^ July 2020. A conscious choice was made to select a period of length 75 days (the number of days between 23rd January 2020 to 6^th^ April 2020). In a similar manner, the most optimal *β* value is selected via minimization of root-mean-square error (RMSE).

To understand the significance of *β* in the paper, we discuss *R*_0_, the basic reproduction number. *R*_0_ is defined as the expected number of cases directly generated by one case in a population where all individuals are susceptible to infection. In particular, *R*_0_ > 1 implies that the infection is spreading in the population and *R*_0_ < 1 indicates that the situation is under control. To obtain an estimate for Singapore’s *R*_0_, we use the different *β* values as obtained using the methodology explained above.

Given

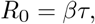

where *R*_0_ = basic reproduction number, *β* = effective contact rate and τ = mean infectious period (time interval during which an individual is infectious), we can calculate the basic reproduction number of COVID-19 in Singapore. By estimating the mean infectious period of COVID-19 τ to be 9 days (Housen et al., 2020; Centers for Disease Control and Prevention (CDC), 2020), we obtain the following range of estimates of *β* and corresponding *R*_0_ in Singapore under the varying degrees of under-estimation of COVID-19 cases in Singapore.

Note that a constant *β* is used across all time periods in the SEIHRD model, whereas we employ a time-varying *β*(*t*) in the SEAIHRD model and reduce the effective contact rate by 5% at selected time points. The time points are selected via an observational study of the different measures employed by the Singapore government to curb the spread of the virus and their perceived corresponding efficacy. In particular, we select the following time points:

– 7^th^ February 2020, when Disease Outbreak Response System Condition (DORSCON) Orange was first declared in Singapore;
– 26^th^ March 2020, when entertainment venues were closed, social gatherings were limited to 10 people and all mass gatherings were cancelled in Singapore.

These 2 time points were chosen as we believe that these 2 events resulted in a substantial decrease in *β*(*t*) under the SEAIHRD model.

From Table 2, all our estimates of *R*_0_ are larger than 1. Coupled along with the exponential increase in Singapore’s COVID-19 cases from mid-April 2020 onward, these 2 pieces of information further support our conjecture that the pandemic was not under control as initially thought.

**Tab. 2:** Fitted dataset used, corresponding *β* and *R*_0_ for SEIHRD and SEAIHRD model

| Fitted dataset | SEIHRD |  | SEAIHRD |  |
| --- | --- | --- | --- | --- |
| | $\beta$ | $R_0$ | $\beta(0)$ | $R_0$ |
| 20% underestimation | 0.35 | 3.13 | 0.40 | 3.57 |
| 40% underestimation | 0.36 | 3.28 | 0.42 | 3.74 |
| 60% underestimation | 0.39 | 3.48 | 0.44 | 3.96 |
| Exponential trend | 0.51 | 4.63 | 0.56 | 5.08 |

### 2.6 Significance of *γ*

Our SEAIHRD model focuses on the differences in robustness and quality of health care offered by different health care systems towards COVID-19 patients, as well as its effect on patient health outcomes. In particular, the pandemic highlighted the preparedness (or lack thereof) of countries’ health care systems to deal with a surge of patients as countries battled successive waves of the virus. Resilient countries displayed their emergency preparedness and continued to maintain a high standard of health care provision, while ill-prepared countries could not adapt and were subsequently crushed by the extra workload, resulting in comparatively higher mortality rates (El Bcheraoui et al., 2020; Mohan, 2021; Chua et al., 2020).

To quantitatively measure the quality of health care provision in Singapore against that of other countries, we look at 3 different metrics, namely the number of physicians per 1,000 people, number of nurses per 1,000 people and number of hospital beds per 1,000 people.

Using The World Bank’s data of the world’s average number of physicians per 1,000 people (1.6) (The World Bank, 2017b), nurses per 1,000 people (3.8) (The World Bank, 2018) and hospital beds per 1,000 people (2.9) (The World Bank, 2017a) as a guideline, we propose the following general formula for *γ* to obtain a non-negative multiplicative constant:

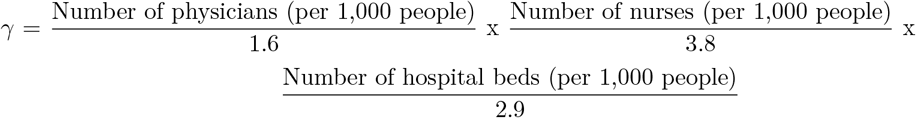

The 3 metrics are chosen as the required information is widely available on public datasets. Furthermore, the metrics are also largely dependent on the state of each country’s health care system, thus allowing us to make a general cross-country comparison of health care provision standards. We made the decision to choose 3 different metrics to minimize the possibility of bias arising from a country’s health care system performing better than the world’s average in only 1 or 2 areas.

We expect our calculation for *γ* to be > 1 in Singapore’s context because Singapore demonstrated a positive response towards the COVID-19 pandemic. Singapore also has the world’s lowest COVID-19 death rate (TODAYonline, 2020), which reflects a more developed health care system as compared to the rest of the world. Using data extracted from the World Bank (The World Bank, 2017a) and MOH (MOH, 2019), our calculation for Singapore’s *γ* equals 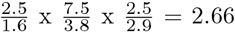. We note that MOH employs the term “doctors” instead of the World Bank’s preferred “physicians”. We do not make a distinction between these 2 groups in the context of our study, considering that the differences in the logical definition of the 2 terms are minor and would not affect our results greatly, if at all. In the absence of complete and up-to-date health care data and information, this value of *γ* serves as a proxy to estimate the strength and resilience of Singapore’s health care system in comparison to that of other countries.

## 3 Simulation Studies

The two epidemiological models (SEIHRD, SEAIHRD) were initialized using deSolve (Soetaert et al., 2020), a R library used for solving differential equations. The initialization values of the parameters for both models are as shown in Table 7 in the Appendix. Due to the lack of clinical data available in the Singapore context, some of the parameter estimates are obtained from other research papers (Singapore Department of Statistics, 2020; Lauer et al., 2020; He et al., 2020; MOH, 2020a; CDC, 2020b; Goh and Chong, 2020; Boëlle et al., 2020). Nonetheless, we are relatively confident of the estimates’ accuracy considering that epidemiological characteristics of COVID-19 is not country-specific and should remain fairly consistent across countries.

### 3.1 Assumptions

To focus on the impact of asymptomatic infections on the total case counts, we have to make several assumptions before we begin our simulation. First, we assumed that *all* asymptomatic patients are automatically unaccounted for in the reported COVID-19 case counts data. Second, we assumed that birth rates and death rates due to other causes are equivalent. Third, based on Singapore’s stringent preventive COVID-19 measures, we made a reasonable assumption that patients who tested positive for COVID-19 are immediately quarantined before being sent to a medical institution to receive treatment. Finally, we assume that all symptomatic patients receive medical treatment in a clinical setting (passing through the *H*(*t*) compartment) before progressing to a health outcome (Recovery / Death). We note that this may be largely true in Singapore or similar urban settings, but such an assumption may be violated in rural areas within large countries such as USA and India, where patients are not able to seek medical treatment in time (Kumar et al., 2020).

### 3.2 Results

## 4 Discussion

At the start of the outbreak, risk of importation of the COVID-19 virus was deemed an existent threat, considering that around 3.4 million people travel from Wuhan to Singapore annually (Koo et al., 2020). Singapore has a population of only 5.7 million (Singapore Department of Statistics, 2020) but boasts one of the highest population densities around the world. For reference, Singapore has nearly 8,000 people per km^2^ more than 200 times as dense as the USA, and 2000 times that of Australia (Ritchie, 2019). This is despite the fact that USA and Australia are more than 13,000 and 10,000 times larger than Singapore respectively in terms of land area. Coupled with one of the highest international visitor arrivals in the world, it resulted in the COVID-19 virus spreading rapidly in Singapore during the early stages of the pandemic.

In this paper, we aimed to showcase the severity of possible under-reporting and reflect the exponential increase in number of COVID-19 cases as a result of the lack of effective intervention methods available during the early months of the outbreak. To do so, we implemented the exponential model, as well as 2 modified SEIR compartmental models; first by incorporating hospitalization dynamics (SEIHRD), before incorporating the presence of asymptomatic cases (SEAIHRD). The 2 epidemiological models were chosen following an extensive review of existing COVID-19 literature and epidemiological models (Arenas et al., 2020; Pazos and Felicioni, 2020), with further modifications made to fit the Singapore context. Using the simulation results from Section 3.2, we can make certain conclusions.

We observe from Figure 6 that with the exception of the 70^th^ quantile of the reported COVID-19 case count growth rate *r*, an exponential model simulation using all other quantiles clearly show the under-estimation of cases in the early stages of the COVID-19 pandemic. However, we note that pandemic simulation is not feasible over long time periods using this model, as shown from the unrealistic trajectory of increase in COVID-19 cases, especially when higher growth rate quantiles from Table 1 are used. Hence, the usage of such a model should be constrained to short-term estimates; for example, to find the date at which the exponential model reaches the cumulative confirmed COVID-19 case count on 6^th^ April 2020 (1,375). From the results in Table 3, we see that the reported COVID-19 case count data is indeed under-estimated when compared against other exponential simulation scenarios. Under the worst-case scenario (90^th^ quantile), we see that the model reaches 1,375 infections on 27^th^ February 2020, a full 39 days earlier as compared to the reported case count data.

**Tab. 3:**
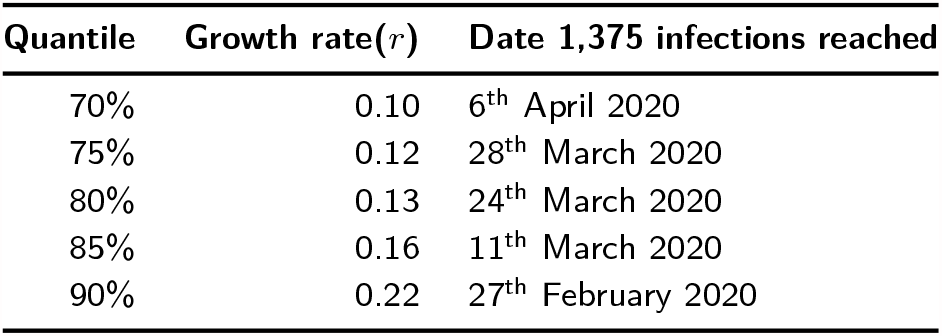
COVID-19 case count growth rate quantiles, corresponding growth rates and date to reach 1,375 infections for exponential model

**Fig. 6:**
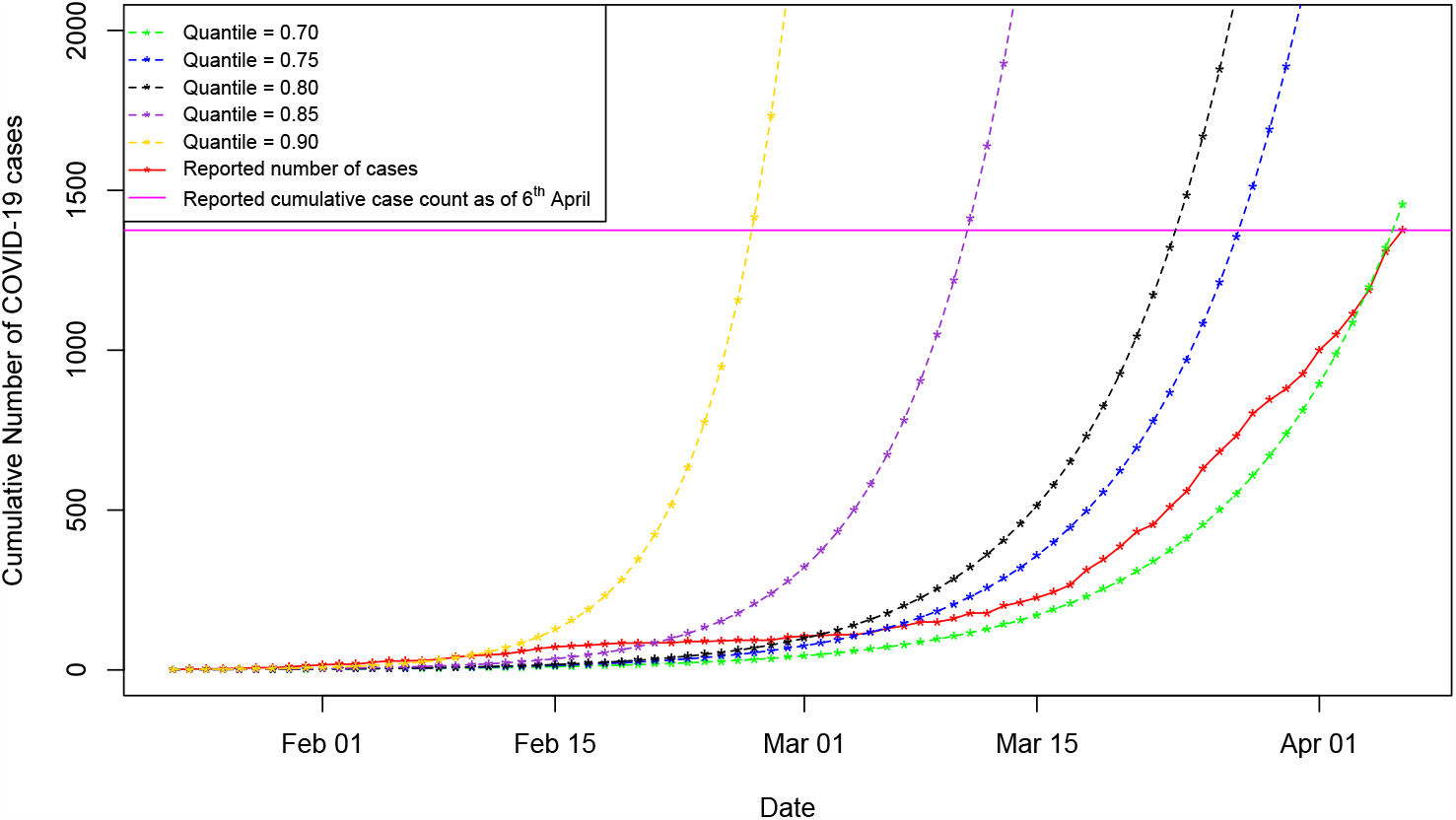
Exponential model simulation results

**Fig. 7:**
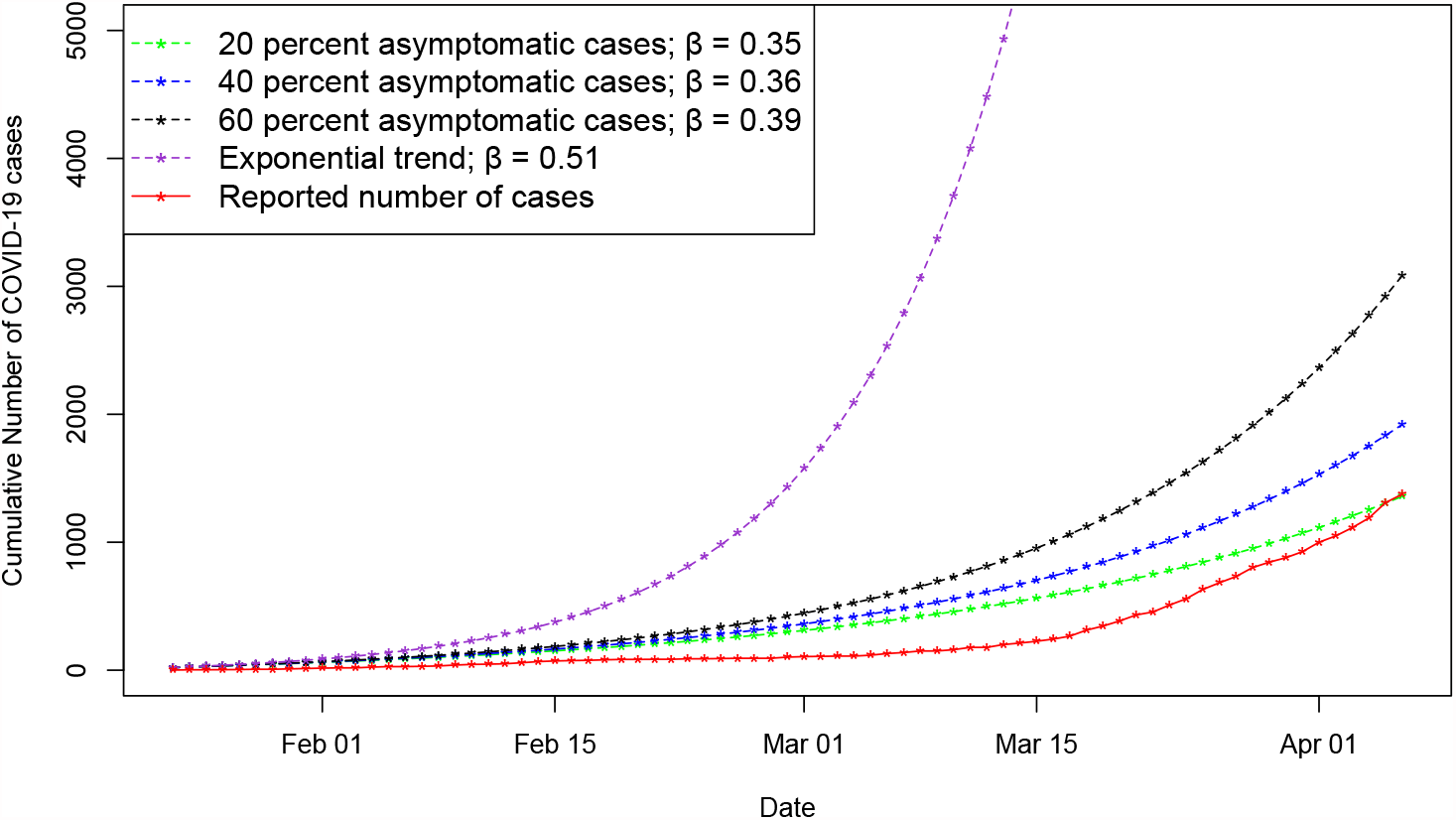
SEIHRD model simulation results

**Fig. 8:**
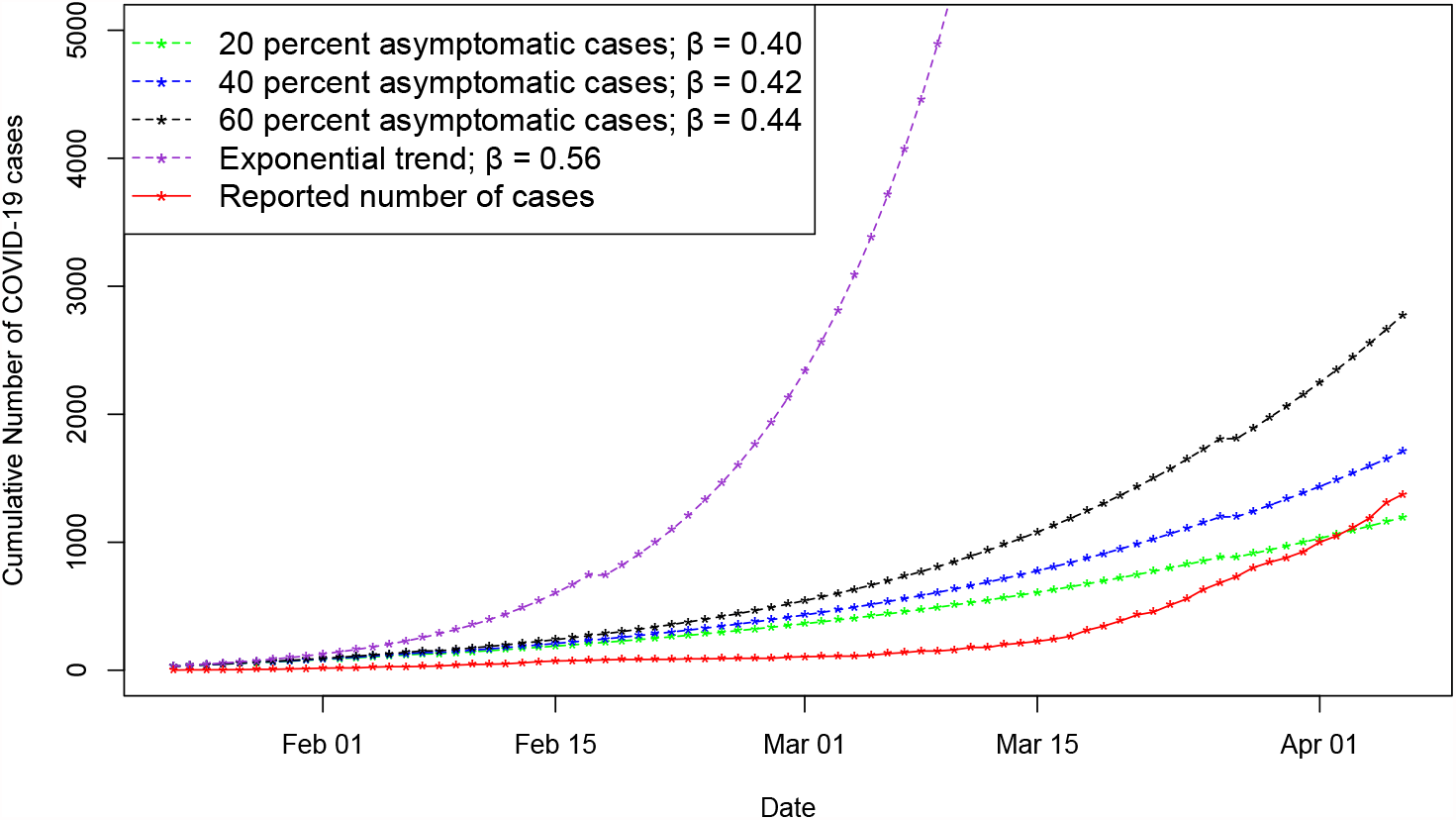
SEAIHRD model simulation results

*β* values corresponding to the 40% (SEIHRD: *β* = 0.36, *R*_0_ = 2.00; SEAIHRD: *β(0)* = 0.42, *R*_0_ = 2.29) and 60% (SEIHRD: *β* = 0.39, *R*_0_ = 2.12; SEAIHRD: *β*(0) = 0.44, *R*_0_ = 2.42) under-estimation in both epidemiological models resulted in an exponential trajectory of COVID-19 cases, greater than the reported case counts. The simulations also reflect the underestimation of the number of COVID-19 cases in the early stages of the pandemic, supporting our findings from the exponential model.

However, the same cannot be said about the *β* values corresponding to the 20% (SEIHRD: *β* = 0.35, *R*_0_= 1.91; SEAIHRD: *β*(0) = 0.40, *R*_0_ = 2.18) under-estimation of cases. Using an under-estimation percentage of 20% gave rise to an over-estimation of cases instead (SEIHRD: -17; SEAIHRD: -181), suggesting that 20% is not a good choice of under-estimation percentage and Singapore’s true COVID-19 case count under-estimation percentage is very likely higher than 20%. At the other end of the spectrum, the *β* values corresponding to the exponential trend (SEIHRD: *β* = 0.51, *R*_0_ = 2.83; SEAIHRD: *β*(0) = 0.56, *R*_0_ = 3.10) result in an unrealistic under-estimation of COVID-19 cases (SEIHRD: 46,820; SEAIHRD: 46,130), considering that Singapore has only reached a cumulative of around 62,000 cases as of June 2021.

Contrary to our expectations at the start of the paper, our SEAIHRD simulations (refer to Table 5) do not consistently display a higher case count across all time periods as compared to our SEIHRD model simulations (refer to Table 4), despite the fact that a higher *β* is used for the SEAIHRD simulations across all under-estimation percentages as compared to the SEIHRD model. This is due to the fact that the SEAIHRD model employs a time-varying *β*(*t*) and the preventive measures taken by the Singapore government are taken into account. Taking into account the differences between the two models’ features (as outlined under Sections 2.3 and 2.4), we conclude that the presence of asymptomatic patients as well as a higher initial effective contact rate in the SEAIHRD model does not outweigh the overall infectivity of a smaller (but constant) effective contact rate in the SEIHRD model. This finding also showcases why Singapore has been so successful at curbing successive waves of the virus; by implementing preventive measures to reduce the overall *β* over time (i.e. decreasing *β*(*t*) as per our approach in the SEIHRD model), this results in a lower case count over an extended period of time.

**Tab. 4:** COVID-19 fitted dataset used, corresponding *β*, extent of under-estimation of cases as of 6th April 2020, corresponding under-estimation % and date for model to reach 1,375 infections under the SEIHRD model.

| Fitted dataset | $\beta$ | Extent of under-estimation | Under-estimation % | Date 1,375 infections reached by model |
| --- | --- | --- | --- | --- |
| 20% underestimation | 0.35 | 17* | 1.22* | NA |
| 40% underestimation | 0.36 | 544 | 39.59 | 30<sup>th</sup> March 2020 |
| 60% underestimation | 0.39 | 1707 | 124.18 | 22<sup>nd</sup> March 2020 |
| Exponential trend | 0.51 | 46820 | 3405.08 | 29<sup>th</sup> February 2020 |

Note: \* denotes an over-estimation of COVID-19 cases by the SEIHRD model

**Tab. 5:** COVID-19 fitted dataset used, corresponding *β*(0), extent of under-estimation of cases as of 6th April 2020, corresponding under-estimation % and date for model to reach 1,375 infections under the SEAIHRD model.

| Fitted dataset | $\beta(0)$ | Extent of under-estimation | Under-estimation % | Date 1,375 infections reached by model |
| --- | --- | --- | --- | --- |
| 20% underestimation | 0.40 | 181* | 13.15* | NA |
| 40% underestimation | 0.42 | 334 | 24.28 | 31<sup>st</sup> March 2020 |
| 60% underestimation | 0.44 | 1400 | 101.82 | 21<sup>st</sup> March 2020 |
| Exponential trend | 0.56 | 46130 | 3354.91 | 25<sup>th</sup> February 2020 |

Note: \* denotes an over-estimation of COVID-19 cases by the SEAIHRD model

Even though the model is capable of demonstrating the under-estimation of COVID-19 cases in Singapore (Griffiths, 2020; Leung, 2020; Illmer, 2020), which is supported by the observation of an exponential increase of confirmed Singapore COVID-19 case counts in later months, we should still remain cautious regarding the predictions at each time point. This is mainly due to our lack of understanding of the virus’ epidemiological characteristics at this point in time, which includes (but not restricted to) the following: (i) incubation period, (ii) extent of contact between susceptible and infectious (asymptomatic / symptomatic / presymptomatic) individuals, (iii) proportion of asymptomatic / symptomatic infections and (iv) transmission rate of asymptomatic / symptomatic individuals.

Any changes to the parameters used in the epidemiological models as a result of our increased understanding of the virus could affect the model’s ability to accurately predict the course of the pandemic. For instance, we do not expect the newer COVID-19 virus variants to possess the same epidemiological characteristics as the original Chinese strain; the parameters would in turn have to be tweaked accordingly to maintain realism and accuracy.

### 4.1 Broader Implications

The three models used are of importance when it comes to pandemic planning and preparedness scenarios. The exponential model could be used as a “worst-case scenario” model (Ghosh et al., 2020) to alert the government of the possible severity of the situation. For example, our best-case simulation scenario of all exponential models (using the 70^th^ quantile of reported COVID-19 case count growth rate) results in an expected case count of 1454, which is close to the confirmed COVID-19 case count at the time point the circuit breaker was announced by the Singapore government. If an assumption was made that the COVID-19 case counts would have increased exponentially (as it has in the following months), more stringent preventive measures could have been taken as soon as the first few confirmed cases were reported in January, thus minimising the ramifications brought about by the virus.

On the other hand, the two other models take longer to build considering the amount of information required about the virus and the state of the country’s health care system. These models are comparatively more complex to develop and execute, but can be used to derive a better understanding of the number of individuals in every compartment at a chosen time point across an extended period of time. Thus, this could help drive better distribution of health care resources such as masks and hand sanitizers and lead to more desirable individual health outcomes.

One can also compare and contrast between the different models to arrive at predictions of higher accuracy based on our understanding of the virus. This would help us better recognise the severity of the situation, and understand the full extent to which case counts are being under-reported as a result of under-testing, which is in turn attributed to our lack of understanding of the infectivity of presymptomatic and asymptomatic cases as discussed in preceding sections.

The COVID-19 case count trajectory under the different models can also be compared against other viral outbreaks in Singapore, such as SARS, H5N1 and H1N1. In the long run, this would aid the government in forecasting and categorising outbreak severity based on different factors such as viral infectivity, presence of and proportion of asymptomatic and presymptomatic cases (if any). This would ultimately support the government’s efforts in developing a robust pandemic mitigation and preparedness plan.

### 4.2 Limitations

Our study has several limitations. First, we can only definitively conclude the underestimation of Singapore’s COVID-19 cases in the early stages of the pandemic based on the exponential increase of confirmed cases in later months, but the degree of underestimation is still unclear due to our lack of understanding of the virus. The degree of under-reporting of cases can only be estimated by conducting a serological surveillance, which relies on the detection of COVID-19 antibodies as a result of a past COVID-19 infection (WHO, 2020). Hence, serological tests are much more reliable and accurate for detecting previous infections in individuals with few or no COVID-19 symptoms (CDC, 2020a). However, Singapore has primarily been adopting the use of polymerase chain reaction (PCR) tests, which involves the extraction of a swab sample from the nose and/or throat from the patient (ICA, 2021). Given the lack of Singapore serological testing results and data, we can only make predictions of the true COVID-19 case counts, subject to the pandemic developing in a similar manner to that of our chosen statistical distributions and accuracy of our parameters based on assumptions made during the development of the models.

Second, the model assumes individuals infected with COVID-19 either recover or die, with no reinfection taking place. New research suggests this might not be true, with countries such as Mexico, Sweden and South Korea all reporting cases of COVID-19 reinfection (Kim, 2020; Richards and Akpan, 2020; de Vrieze, 2020). Third, we assumed in our model that patients who are asymptomatic, or showing little to no symptoms eventually transition to the (R)ecovered compartment in the epidemiological model. However, as discussed in Section 2.4, asymptomatic patients may also suffer from other symptoms in the long run. This is supported by research showing that 3 in every 4 COVID-19 patients suffer from at least one symptom six months after infection (Khalik, 2021). The severity of these symptoms, and their impact on future COVID-19 cases / related deaths in the long run are still unknown at this point in time.

## 5 Conclusion

Pandemic planning is crucial at the early stages of a virus to avert a nationwide outbreak. In this study, we investigated the impact of presymptomatic and asymptomatic infections on the total number of COVID-19 cases, and estimated the true basic reproduction number *R*_0_ in the early stages of the pandemic. Using different epidemiological models, we find that under the assumption of different asymptomatic proportions and effective contact rates, the trajectory of COVID-19 cases increased exponentially higher than reported figures. Our experience and lessons learnt will contribute to pandemic preparedness for any future outbreaks and mitigation, minimising any negative societal and economic impacts.

## Data Availability

The data that support the findings of this study are openly available in **Singapore: Coronavirus Pandemic Country Profile** at https://ourworldindata.org/coronavirus/country/singapore.

## Appendix

**Tab. 6:**
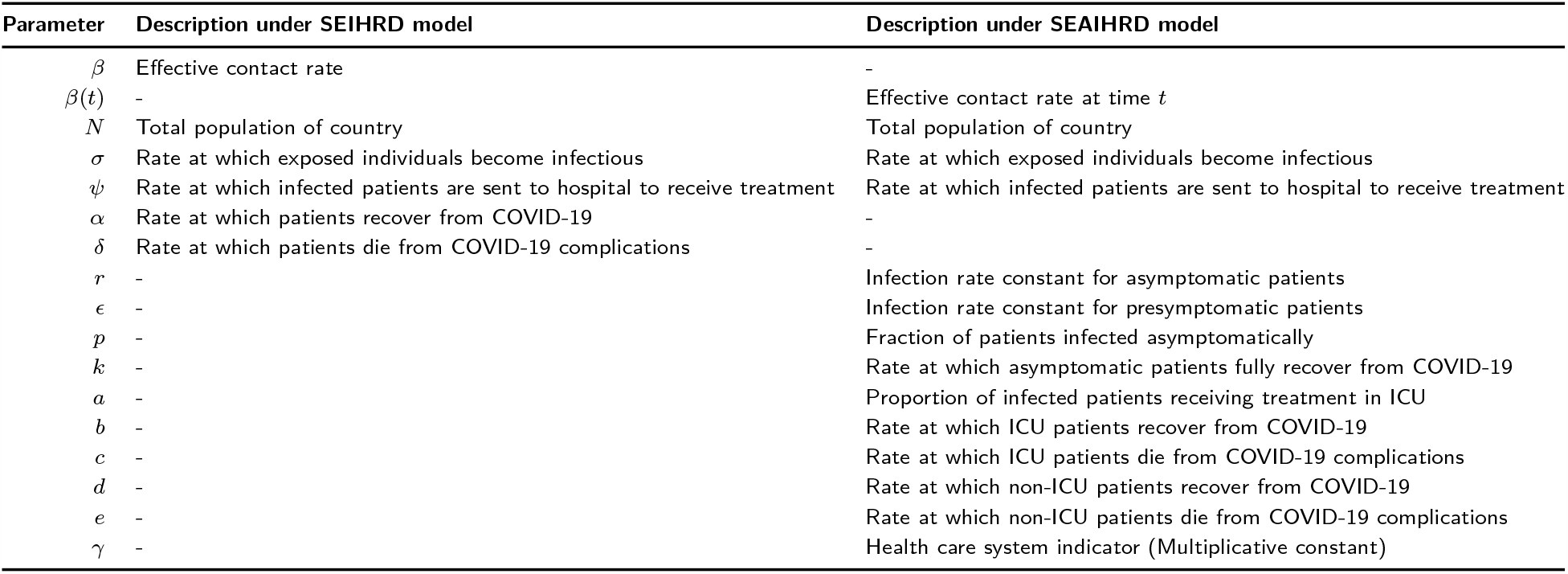
Description of parameters used in SEIHRD and SEAIHRD model

**Tab. 7:**
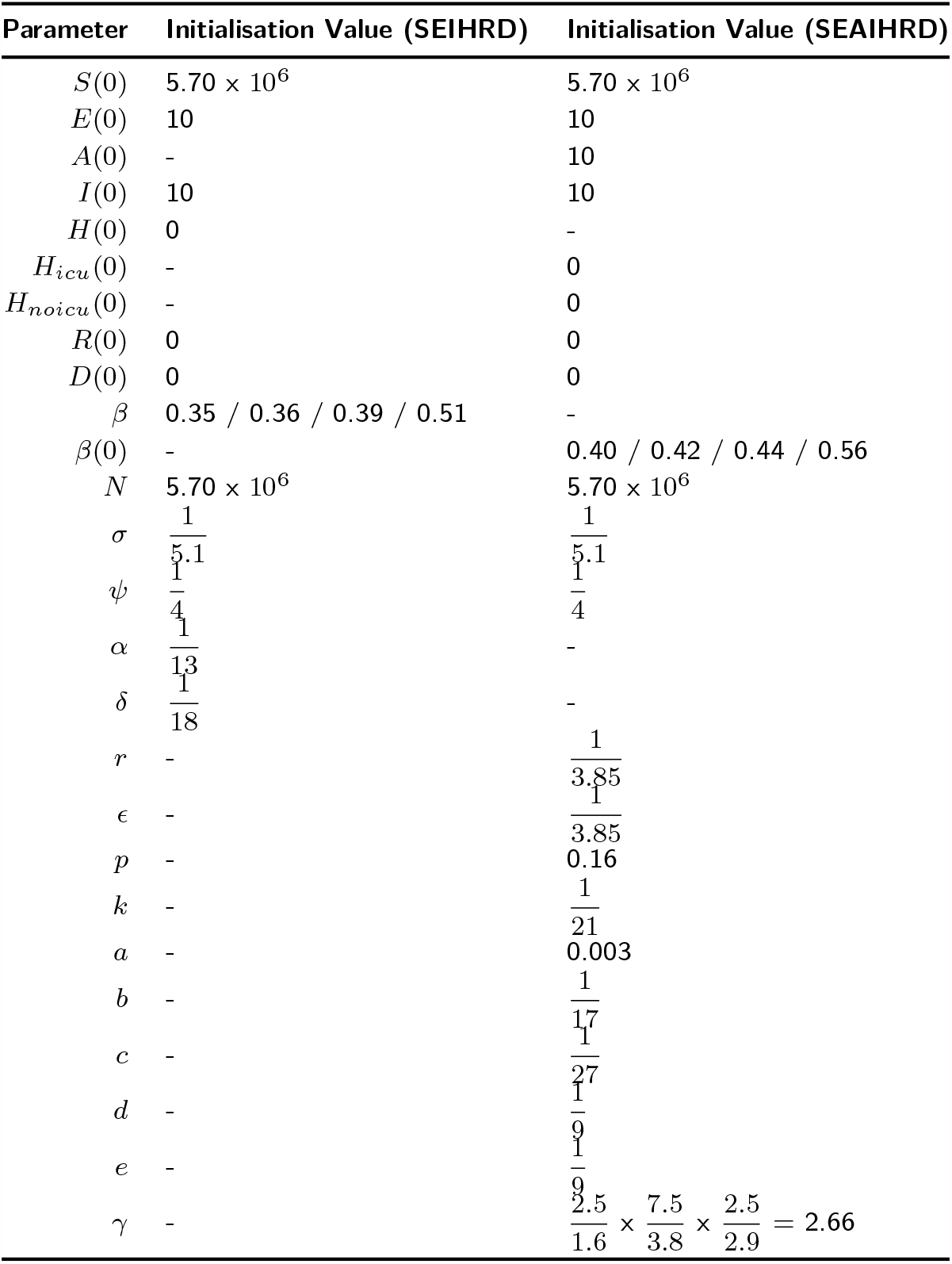
Parameters and Initialization Values used in Singapore’s SEIHRD / SEAIHRD model

